# Clustering and Healthcare Costs With Multiple Chronic Conditions in a US Cross-Sectional Study

**DOI:** 10.1101/2020.08.24.20179184

**Authors:** C Hajat, Y Siegal, Amalia Adler-Waxman

**Affiliations:** Public Health Institute, UAE University; Deloitte Consulting LLP; Teva Pharmaceuticals Industries Ltd

**Keywords:** Multiple Chronic Conditions, Multimorbidity, Healthcare Costs, Clusters, Cardiovascular

## Abstract

**Objective:** To investigate healthcare costs and contributors to costs for multiple chronic conditions (MCCs), common clusters of conditions and their impact on cost and utilisation.

**Methods:** This was a cross-sectional analysis of US financial claims data representative of the US population, including Medicare, Medicaid and Commercial insurance claims in 2015. Outcome measures included healthcare costs and contributors; ranking of clusters of conditions according to frequency, strength of association and unsupervised (k-means) analysis; the impact of clustering on costs and contributors to costs.

**Results:** Of 1,878,951 patients, 931,045(49.6%) had MCCs, 56.5% weighted to the US population. Mean age was 53.0 years(SD16.7); 393,121(42.20%) were male. Mean annual healthcare spending was $12,601, ranging from $4,385 (2 conditions) to $33,874 (11 conditions), with spending increasing by 22-fold for inpatient services, 6-fold for outpatient services, 4.5-fold for generic drugs and 4.2-fold for branded drugs.

Cluster ranking using the 3 methodologies yielded similar results: highest ranked clusters included metabolic syndrome(12.2% of US insured patients), age related diseases(7.7%), renal failure(5.6%), respiratory disorders(4.5%), cardiovascular disease(CVD)(4.3%), cancers(4.1-4.3%), mental health-related clusters(1.0-1.5%) and HIV/AIDS(0.2%).

Highest spending was in HIV/AIDS clusters ($48,293), mental health-related clusters ($38,952-$40,637), renal disease ($38,551) and CVD ($37,155); with 89.9% of spending on outpatient and inpatient care combined, and 10.1% on medication.

**Conclusion and Relevance:** Over 57% of insured patients in the US may have MCCs. MCC Clustering is frequent and is associated with healthcare utilisation. The findings favour health system redesign towards a multiple condition approach for clusters of chronic conditions, alongside other cost-containment measures for MCCs.

**Highlights:** *What is already known about the topic?:* Despite one in three adults suffering from more than one chronic condition, little is known about the burden from MCCs. Some studies to date suggest markedly different disease, cost and societal burdens. Furthermore, certain conditions cluster together more frequently, however, no studies have reported on the impact that clusters have on the healthcare cost burden from MCCs in a comprehensive manner. Recent consensus statements have called for a specific focus on multiple chronic conditions.

*What does the paper add to existing knowledge?:* This study is one of the most comprehensive studies investigating contributors to costs in terms of number of patients included, representativeness of the US population and inclusion of the full range of chronic conditions.

*What insights does the paper provide for informing healthcare-related decision making?:* Of US insured patients, over 57% may have multiple chronic conditions. HIV/AIDS was the costliest cluster followed by clusters of mental and behavioural disorders, renal failure and CVD. Outpatient and inpatient services account for roughly 90% of health spending and medication for 10%. Health service utilisation varies by number and clusters of conditions, with potential overutilization of specialist services and underutilisation of primary care and psychiatric services.

## Introduction

The increasing global burden of non-communicable diseases (NCDs), accounting for three in five of global deaths [1], has long been recognised as a global priority. Less attention has been given to the issue of multiple chronic conditions (MCCs), also termed multi-morbidity, despite one in three adults suffering from more than one chronic condition [2].

Evidence from a handful of studies reporting on the burden from MCCs to date suggest that this phenomenon results in markedly different disease, cost and personal burdens. Most studies have asserted a positive association between MCC and healthcare expenditures [3], some reporting a doubling in costs with each subsequent condition [4,5]. Studies suggest more complex inpatient and outpatient care utilisation and use of more prescription medications [6-9].

Certain conditions cluster together more frequently such as stroke and Alzheimer’s disease, and communicable conditions such as TB and HIV/AIDS with diabetes and CVD, respectively [10,11]. However, no studies have reported on the impact that clusters have on the healthcare cost burden from MCCs in a comprehensive manner.

Recent articles have called for a specific focus on multiple chronic conditions [10-12] and the Academy of Medical Sciences in the UK identified clustering of disease as a priority research area [13]. The objective of this article is to quantify healthcare spending in patients with MCCs using a comprehensive set of chronic conditions, identify the most important clusters and identify the key contributors to costs for MCC patients.

## Methods

The primary research questions were:

1. What are the healthcare costs and contributors to costs for patients with multiple chronic conditions?
2. What are the most important clusters of chronic conditions?
3. How does clustering of chronic conditions impact cost and contributors to costs?

We used a random sample of MarketScan® claims-based data [14] for data year 2015 including the Commercial Claims and Encounters (CCAE) Database, the Medicare Supplemental Database, and the Multi-State Medicaid Database. The MarketScan^®^ databases contain de-identified, patient-level health episode claims information including inpatient services, outpatient services and outpatient prescription drugs. The CCAE database covers patients with commercial insurance while the Medicare Supplemental Database and the Multi-State Medicaid Databases covers patients submitting claims through the public programmes of Medicare and Medicaid, respectively [15]. The MarketScan database utilises standard international coding and the International Classification of Disease [16] was used to assign diagnoses.

Patients with two or more chronic conditions from the Agency for Healthcare Research and Quality (AHRQ) list of 69 chronic conditions which relies on ICD9 and ICD10 coding systems [17] were included. Patients with more than 11 conditions were omitted due to inadequate sub-group numbers. We employed a two-step process to identify and select clusters for further analyses and applied different methodologies due to the absence of one agreed methodology to identify clusters of disease.

Step 1 was to identify the top 25 ranked condition pairs according to the attributes of chronic conditions that are most relevant to clusters, those of frequency and strength of association. An unsupervised clustering analysis (k-means clustering method) was conducted to identify clustering without constraining to condition pairs. K-means is an unsupervised learning method without reliance on a “dependent” variable. K-means clustering optimizes the within group sum of squares i.e. it assigns observations to a cluster based on how close (Euclidean distance) it is to the cluster centroid. Each patient was assigned to a disease cluster based on Euclidean distance of their disease vectors to the cluster centroids [18]. The clusters are reported in the results tables characterised by the base conditions which strongly associate with the cluster (i.e., are present in ≤95% of patients in that cluster), as well other conditions more moderately associated with the cluster (present in 20-95% of patients in that cluster).

Step 2 was to select 10 clusters to investigate further with meaningful results. There were selected according to ranking and representative of a range of conditions and global distribution. For example, clustering around HIV/AIDS was included for further analyses as the only cluster of a chronic communicable condition ranking in the top 25 and meeting the above conditions.

Contributors to costs that were investigated included: age, sex, number of conditions, clustering of conditions, costs from inpatient, outpatient and other services, branded and generic medications, and of medical services: specialty procedures and diagnostics, primary care, emergency visits and psychiatric services. Definitions of healthcare spending are detailed in the technical supplement.

To make the results representative of the US insured population distribution, scaling factors were used to weight samples from each coverage type to the corresponding US adult (18+) insured population from year 2015 (including 134.1 million CCAE, 43.3 million Medicare and 29.6 million Medicaid insured population) [19].

### Statistical analysis

Analyses were conducted using the software package R, version 3.4.0 (2017-04-21). Anova testing was used to determine the statistical significance of differences in costs (for each sub-category e.g., outpatient costs, inpatient costs, medication costs) between clusters (at the 95% significance level) adjusted for age and sex. Cancers were stratified into male and female types and adjusted for age only. All results were reported after weighting to the US population other than the crude figures used for reporting the study sample patient characteristics (table 1).

**Table 1.** Characteristics of Patients Included.

| Characteristic |  |
| --- | --- |
| Number with multiple chronic conditions | 931,045 (49.6%) |
| Age years: mean(SD) | 53.04 (16.66) |
| Male n(%) | 393,121 (42.20%) |
| Health Coverage: n(%) |  |
| Commercial (CCAE) | 476,879 (51.23%) |
| Medicaid | 270,092 (29.00%) |
| Medicare | 184,074 (19.77%) |
| *Number of chronic conditions:* |  |
| 2 n(%) | 201,255 (21.60%) |
| 3 n(%) | 167,651 (18.00%) |
| 4 n(%) | 134,020 (14.40%) |
| 5 n(%) | 103,575 (11.10%) |
| 6 n(%) | 78,844 (8.47%) |
| 7 n(%) | 60,056 (6.45%) |
| 8 n(%) | 45,449 (4.88%) |
| 9 n(%) | 34,649 (3.72%) |
| 10 n(%) | 26,078 (2.80%) |
| 11 n(%) | 20,036 (2.15%) |
| Annual Healthcare spending US\$: mean(SD) | |
| *Overall* | 12,601 (36,329) |
| *By type of coverage:* |  |
| Commercial (CCAE) | 10,571 (28,352) |
| Medicaid | 11,729 (38,518) |
| Medicare | 19,139 (48,596) |

As coverage type criteria include age, comparisons between coverage types were not adjusted by demographic factors. Other patient factors such as socio-economic status and ethnicity were not available.

### Patient and Public Involvement

This work forms part of a broader study in which focus group work was conducted on patients with MCCs to elicit their main struggles and concerns and the terminology used in discussing their conditions. These qualitative insights were published in a white paper [11] and the current study aims to answer the quantitative research questions.

## Results

Table 1 reports characteristics of the study sample. The data comprised 51.2% of CCAE, 29.0% Medicaid and 19.8% Medicare patients. Of an initial sample of 1,878,951 subjects, at-least 2 chronic conditions were found in 931,045 (49.6%); when scaled to the US population, this would equate to 56.5%. Mean age was 53.0 years (SD16.7); 393,121 (42.20%) were male and 871,613 with between 2-11 conditions were further analysed. Mean annual healthcare spending was $12,601, varying between the types of insurance at $10,571 for Medicaid, $11,730 for Medicare and $19,139 for CCAE (p<0.001).

Figure 1 shows mean healthcare spending per patient and contributors to costs, according to the number of chronic conditions. Average healthcare spending per annum for those with two conditions was $4,385, increasing eight-fold to $33,874 for those with 11 or more conditions.

**Figure 1.**
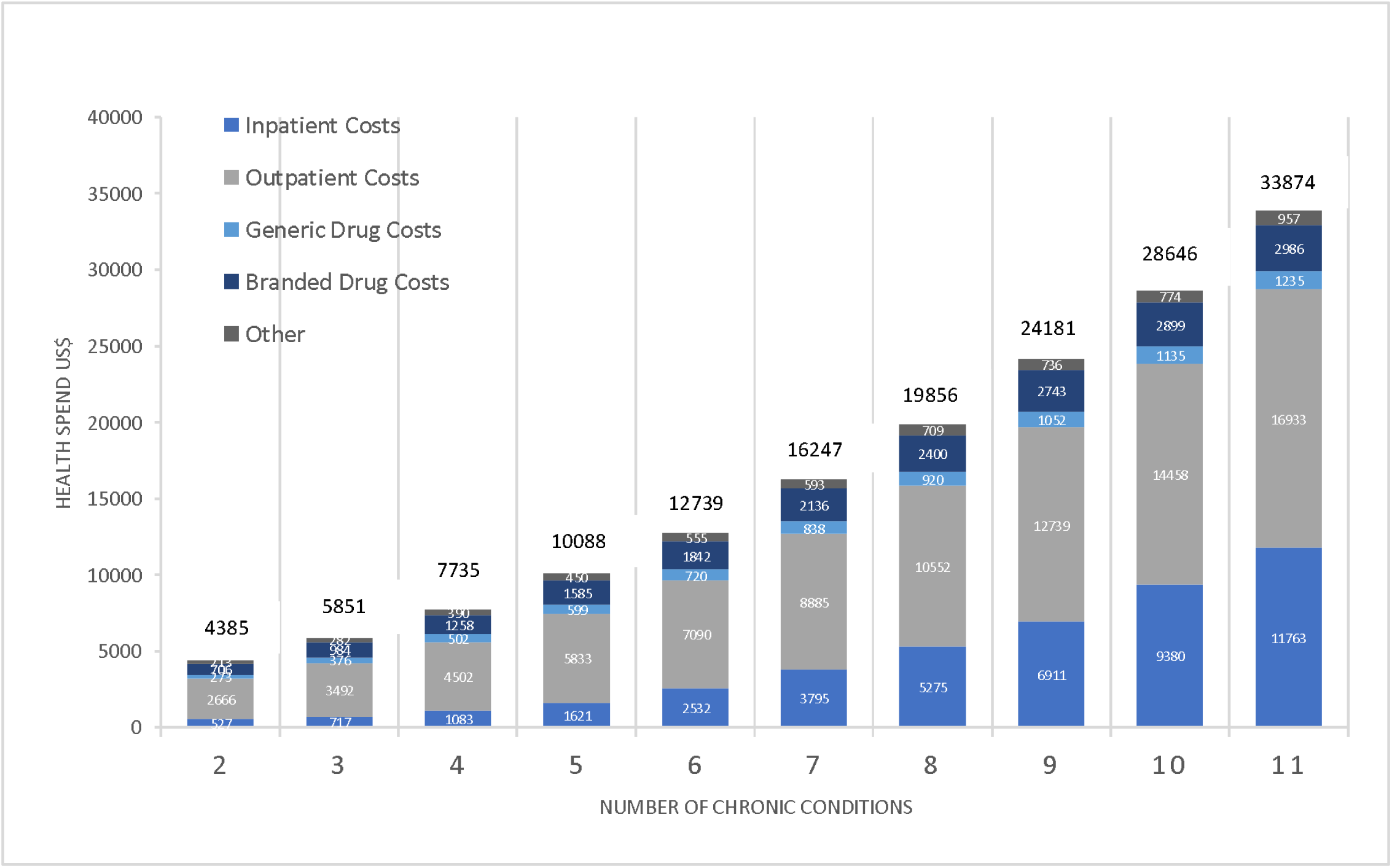
Total Average Health Spend and Cost Drivers by Number of Chronic Conditions. Costs are weighted to the US population and are reported in US$. Comparison between costs by number of conditions was statistically significant overall (p<0.001), for inpatient (p<0.001), outpatient (p<0.001), branded drugs (p<0.001) and generic drugs (p<0.001).

Whilst the absolute values increased, the percentage change was non-linear, showing a consistently gradual decrement from 33% (shift from two to three conditions) to 18% (shift from 10 to 11 conditions). Inpatient costs accounted for the greatest shift in both absolute and relative amounts. The relative increase in healthcare spending between patients with 2 and 11 conditions was 22-fold for inpatient costs, 6-fold for outpatient costs, 4.5-fold for generic drugs and 4.2-fold for branded drugs.

Further analysis of the purely medical contributors to costs for MCC patients showed that between having 2 and 11 conditions, healthcare spending increased by 24-fold for inpatient services, 8-fold for speciality procedure and diagnostics, 6-fold for emergency visits and 4-fold for both primary care services and psychiatric services.

Table 1 here and technical supplement table 1 show that average spending per annum was highest for Medicare at $19,129 and considerably lower for both Medicaid at $11,783 and CCAE at $10,572 (p<0.001). Younger patients on Medicaid had higher spending on male cancers, renal failure and HIV/AIDS whilst older patients on Medicare had higher spending for clusters associated with mental health problems, age-related disease and respiratory problems. Table 1 shows that the overall average spending is higher in Medicare than in CCAE, however the technical supplement table 1 shows that when number of conditions is held fixed, Medicare spending is lower than CCAE due to typically lower reimbursement in Medicare compared with CCAE for any given condition.

Table 2 here shows the ten clusters included in further analyses; technical supplement tables 2, 3 and 4 show the list of top 25 clusters ranked by frequency, strength of association and unstructured k-means clustering from which the ten clusters were derived. Among these were metabolic syndrome, present in 12.2% of the insured population and predominantly including hypertension, high cholesterol and diabetes mellitus. Renal failure (present in 5.6%) and cancers (present in 4.1-4.3%) clustered with hypertension, high cholesterol, diabetes and other neurological disorders, amongst other conditions. Clusters of age-related diseases (present in 7.7%) included osteoarthritis, hypertension and high cholesterol, and cardiovascular disease (CVD), present in 4.3%, included hypertension, coronary artery disease (CAD), high cholesterol and cardiomyopathy. Respiratory disorders (present in 4.5%) including chronic obstructive pulmonary disease (COPD) and allergy, clustered with hypertension, high cholesterol, peptic ulcer disease (excluding bleed), depression and other neurological disorders. Mental health disorder clusters, including depression and anxiety, were present in 1.0-1.5% and clustered with chronic pain, hypertension, high cholesterol, diabetes, other neurological disorders and alcohol abuse. The HIV/AIDS cluster was present in 0.2% of the insured population and occurred with hypertension, high cholesterol, alcohol abuse, depression and weight loss.

**Table 2.**
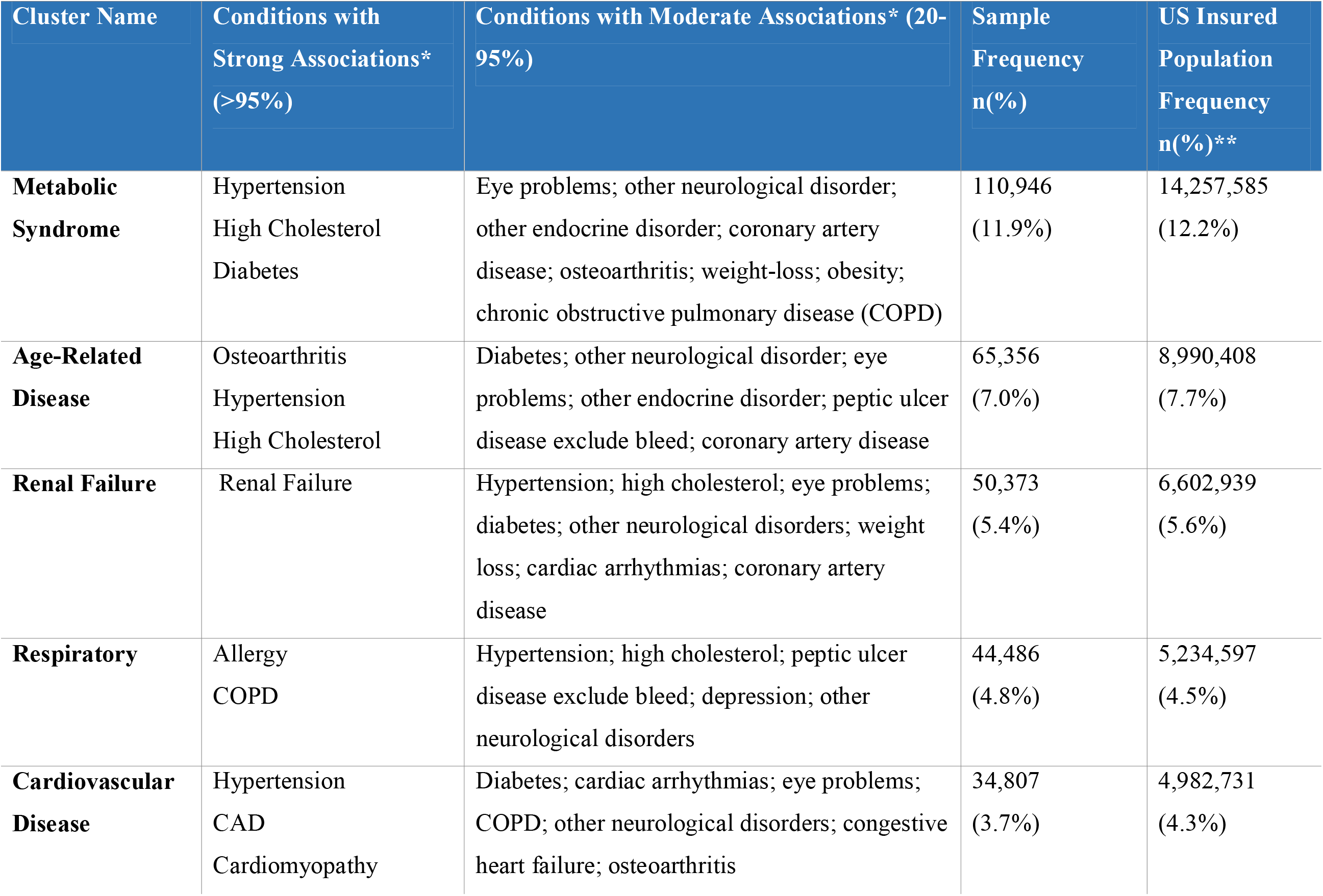
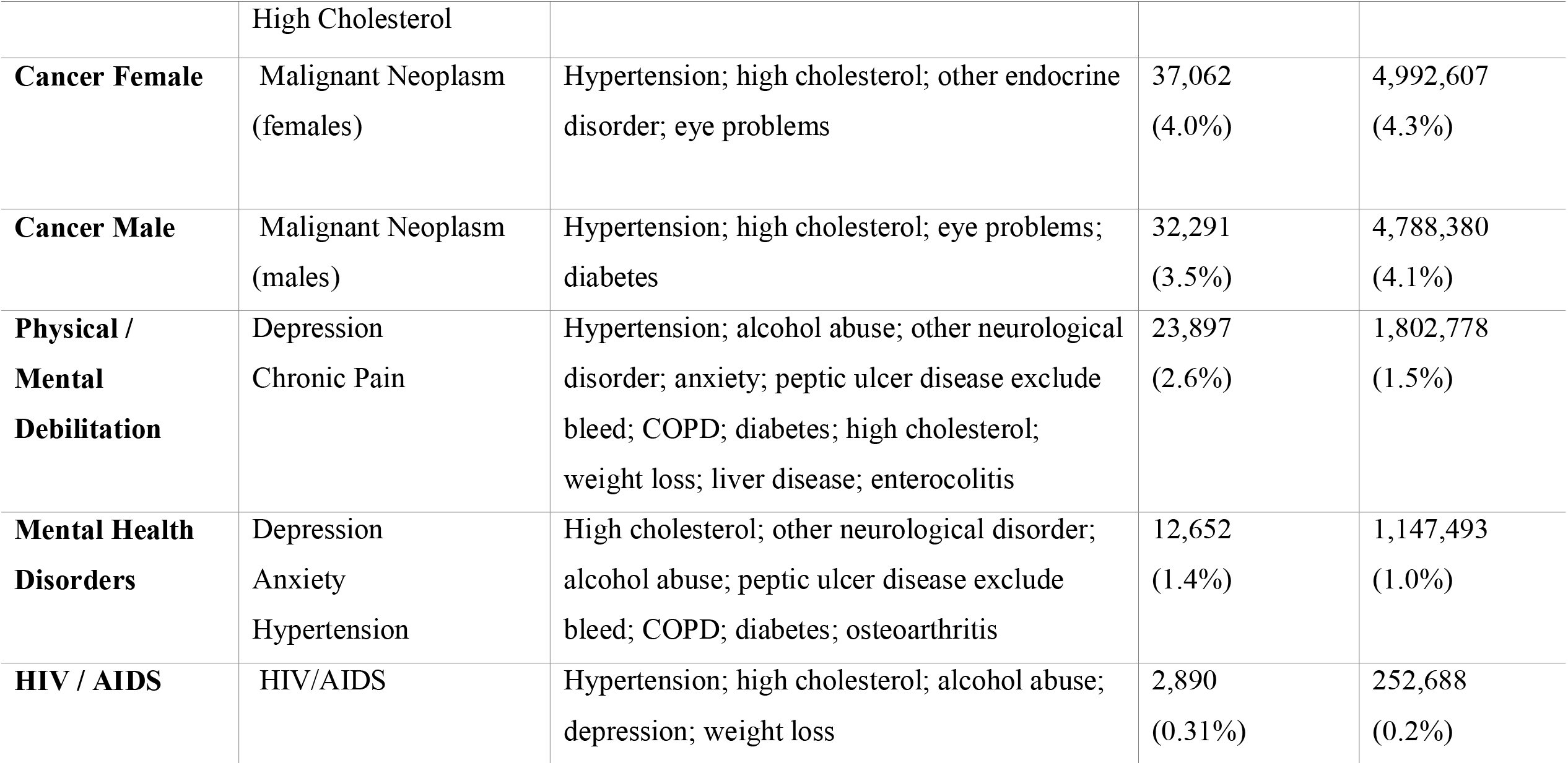
Top Clusters of Chronic Conditions. *The second column reports the main chronic conditions that were present in over 95% of patients falling within that cluster; column three reports conditions that were present in 20-95% of patients within that cluster. **Weighted and extrapolated to the US insured population

| Cluster Name | Conditions with Strong Associations* (>95%) | Conditions with Moderate Associations* (20-95%) | Sample Frequency<br>n(%) | US Insured Population Frequency<br>n(%)** |
| --- | --- | --- | --- | --- |
| **Metabolic Syndrome** | Hypertension<br>High Cholesterol<br>Diabetes | Eye problems; other neurological disorder; other endocrine disorder; coronary artery disease; osteoarthritis; weight-loss; obesity; chronic obstructive pulmonary disease (COPD) | 110,946<br>(11.9%) | 14,257,585<br>(12.2%) |
| **Age-Related Disease** | Osteoarthritis<br>Hypertension<br>High Cholesterol | Diabetes; other neurological disorder; eye problems; other endocrine disorder; peptic ulcer disease exclude bleed; coronary artery disease | 65,356<br>(7.0%) | 8,990,408<br>(7.7%) |
| **Renal Failure** | Renal Failure | Hypertension; high cholesterol; eye problems; diabetes; other neurological disorders; weight loss; cardiac arrhythmias; coronary artery disease | 50,373<br>(5.4%) | 6,602,939<br>(5.6%) |
| **Respiratory** | Allergy<br>COPD | Hypertension; high cholesterol; peptic ulcer disease exclude bleed; depression; other neurological disorders | 44,486<br>(4.8%) | 5,234,597<br>(4.5%) |
| **Cardiovascular Disease** | Hypertension<br>CAD<br>Cardiomyopathy | Diabetes; cardiac arrhythmias; eye problems; COPD; other neurological disorders; congestive heart failure; osteoarthritis | 34,807<br>(3.7%) | 4,982,731<br>(4.3%) |
|  | High Cholesterol |  |  |  |
| **Cancer Female** | Malignant Neoplasm<br>(females) | Hypertension; high cholesterol; other endocrine<br>disorder; eye problems | 37,062<br>(4.0%) | 4,992,607<br>(4.3%) |
| **Cancer Male** | Malignant Neoplasm<br>(males) | Hypertension; high cholesterol; eye problems;<br>diabetes | 32,291<br>(3.5%) | 4,788,380<br>(4.1%) |
| **Physical /**<br>**Mental**<br>**Debilitation** | Depression<br>Chronic Pain | Hypertension; alcohol abuse; other neurological<br>disorder; anxiety; peptic ulcer disease exclude<br>bleed; COPD; diabetes; high cholesterol;<br>weight loss; liver disease; enterocolitis | 23,897<br>(2.6%) | 1,802,778<br>(1.5%) |
| **Mental Health**<br>**Disorders** | Depression<br>Anxiety<br>Hypertension | High cholesterol; other neurological disorder;<br>alcohol abuse; peptic ulcer disease exclude<br>bleed; COPD; diabetes; osteoarthritis | 12,652<br>(1.4%) | 1,147,493<br>(1.0%) |
| **HIV / AIDS** | HIV/AIDS | Hypertension; high cholesterol; alcohol abuse;<br>depression; weight loss | 2,890<br>(0.31%) | 252,688<br>(0.2%) |

The three different methods of identifying clusters reported in detail in technical supplement tables 2, 3 and 4 yielded the same conditions for the majority of the top 25 clusters with clustering of high cholesterol with hypertension, diabetes with hypertension, high cholesterol with other endocrine disorders and osteoarthritis with hypertension ranking highly. Of the top 25 condition pairs by strength of association, only three individual conditions did not also appear in the top 25 by frequency, namely coagulopathy, blood loss anaemia and cystic fibrosis.

Figure 2 shows the average annual healthcare spending and contributors to costs by clusters. The highest spending was for patients with the HIV/AIDS cluster at $48,293 per patient per annum, predominantly driven by the cost of branded drugs costing on average $21866 (45.3%) of their total spending. Subsequent ranks by spending included the clusters of mental/physical debilitation at $40,637, mental health disorders at $38,952, renal disease at $38,551 and CVD at $37,155, with 45-50% of the healthcare spending on outpatient care, 40-45% on inpatient care and 10% on medication costs. In addition to the clusters detailed here, other clusters that were associated with “other neurological disorders” accounted for higher healthcare spending in general.

**Figure 2.**
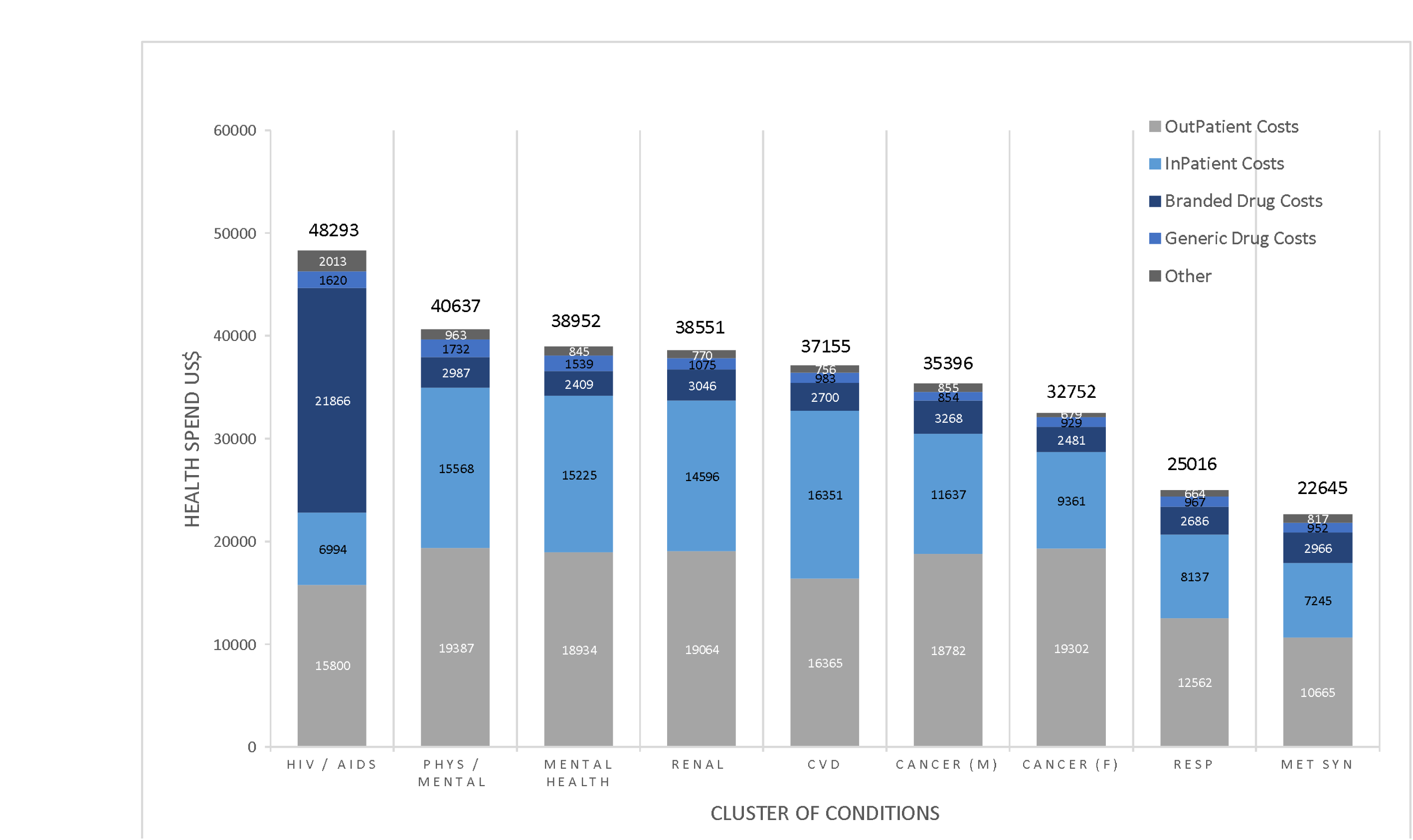
Total Average Health Spend and Cost Drivers by Cluster of Conditions. Costs are weighted to the US population and reported in US$. Comparisons for cost between clusters of conditions were statistically significant overall (p<0.001), for inpatient (p<0.001), outpatient (p<0.001), branded drugs (p<0.001) and generic drugs (p<0.001).

Health Service Utilisation was dependent on both the number and cluster of conditions. The proportion of total health spending on inpatient costs increased with each additional condition from $515(20.7%) for 2 conditions to $12,292 (47.8%) with 11 conditions. The proportion of total health spending on primary care services decreased with each additional condition from $362 (14.6%) for 2 conditions to 1,490 (5.8%) for 11 conditions. Figure 3 shows the pattern of utilisation of health services by cluster. Inpatient services accounted for over half of medical health spending for many of the clusters, followed closely by spending on specialty procedures and diagnostics. Spending on psychiatric services was low for all clusters, ranging from $58 (0.2%) in metabolic syndrome to $210 (1.4%) in the HIV/AIDS cluster and $418 (1.5%) in the mental health disorder cluster.

**Figure 3.**
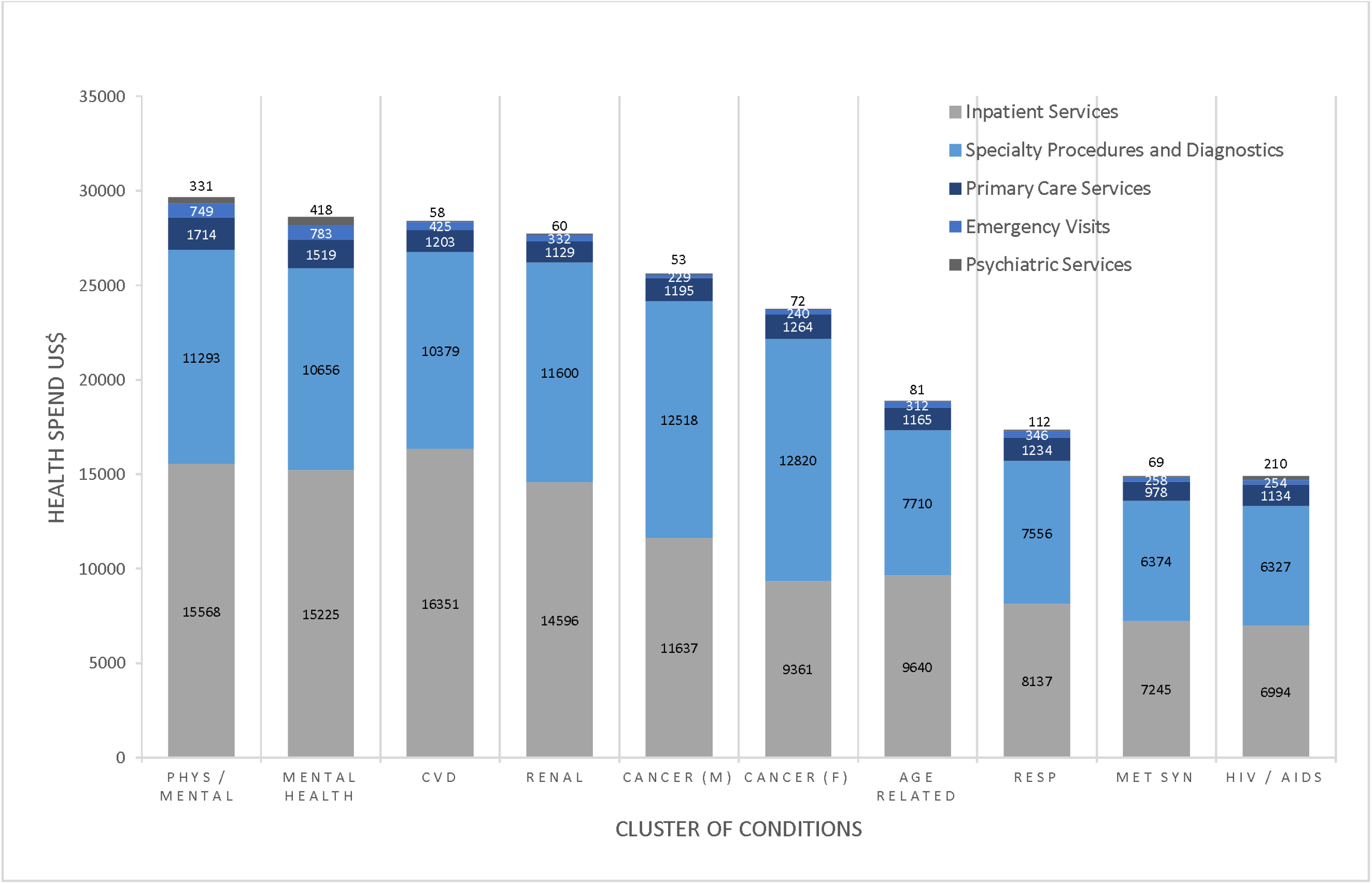
Health Service Utilisation in terms of cost, reported by Cluster. Costs are weighted to the US population and are reported in US$. Comparisons for cost between clusters of conditions were statistically significant for inpatient (p<0.001), specialty procedure (p<0.001), primary care (p<0.001), emergency care (p=0.003) and psychiatric care (p<0.001).

Healthcare spending by clusters stratified by age group followed predicted patterns of higher spending for older age groups in the clusters of metabolic syndrome, mental health disorders, physical and mental debilitation, HIV/AIDS, respiratory and age-related conditions. However other clusters such as CVD, female cancers and renal failure varied less between age groups. Mean healthcare spending for CVD was $34,485 for 18-44 year olds and $38,175 for those aged 85 and over, with similar outpatient, inpatient and generic drug costs. Spending on branded drugs accounted for a large proportion of the variance ($3,121 and $2,703 in ages 45-64 & 65-84, respectively compared with $1932 and $1,545 in ages 18-44 and >85, respectively; p<0.001). Male cancers are costlier in younger age groups ($42,118 for ages 18-44 compared with $33,206 for age 85; p<0.001) due to the specific types of cancer seen in younger men.

## Discussion

This study represents one of the most comprehensive studies to date to investigate contributors to costs in terms of number of patients included, representativeness of the US population and because it investigates the full range of, rather than selected, chronic conditions.

Key findings include:

1. In this large US insured patient sample, 50% had multiple chronic conditions.
2. Costs of healthcare spending increase non-linearly with each subsequent condition. Our study shows a relatively smaller increment in healthcare spending compared with previous smaller published studies [10], and that the relative increase tapers off with each subsequent chronic condition.
3. Overall, inpatient costs accounted for the highest increase with each subsequent condition, with a 24-fold increase between patients with 2 and 11 conditions, with considerable variation according to the ‘cluster’ of conditions.
4. The most important clusters identified were, in decreasing frequency, metabolic syndrome conditions (present in 12%), chronic renal failure (8%), age related diseases (7%), respiratory disorders (4.5%), CVD (4%), cancers (4%) and mental and behavioural disorders (1-1.5%). In addition, the HIV/AIDS cluster, of particular interest to LMIC settings, was present in 0.2%.
5. Conditions occurring in over 95% of patients with these clusters included hypertension, high cholesterol, diabetes, CVD, depression, anxiety, chronic pain, osteoarthritis, allergy, COPD, male and female cancers, chronic renal failure and HIV/AIDS.
6. HIV/AIDS was the costliest cluster due to 45% of total spending on branded drugs. This was followed by clusters of mental and behavioural disorders which were second and third most costly and, renal failure and CVD at fourth and fifth most costly, with outpatient and inpatient services accounting for roughly 90% of health spending.
7. Health service utilisation varied by number of condition and clusters, with possible overutilization of specialist services and underutilisation of primary care and psychiatric services in those with additional chronic conditions and certain clusters.

### Frequency Rates

Our sample of one year of hospital episode data showed that exactly a half (56.5% scaled to the US population) of patients had at-least 2 chronic conditions. However, as this is a patient sample it does not seek to represent the US population prevalence rate which would be expected to be lower due to the presence of non-healthcare seeking adults. Nonetheless, the rates are within range of previously reported rates.

Prevalence estimates for MCC are highly heterogeneous with methodological differences such as age, the number of chronic conditions included and whether the outcomes are self-reported or verified leading to estimates that may vary up to threefold. Prevalence estimates for MCC range from 25.5% in the US (for 10 chronic conditions), increasing in the US to 50% for ages 45 to 65 and 81% for ages over 65 years [20], 16% in the UK (for 17 chronic conditions) to 58% (for 114 chronic conditions) [21], 45% in China to 71% in Russia in those aged over 50 [22] and just 9.4% in India [23].

### Costs and number of conditions

A few previous studies reported healthcare spending doubling with each additional chronic condition [4,5] whilst others reported smaller increments [3]. Our study shows a modest increase in healthcare spending and that the relative increase tapers off with each subsequent chronic condition. As our study is much larger and includes the full spectrum of chronic conditions, it is likely to be more representative of the overall status for chronic conditions.

### Clusters

This is one of the most comprehensive studies to date that identifies and quantifies clustering between the full range of chronic conditions. Conditions may cluster together by virtue of independently high prevalence rates, shared risk factors and disease pathways or due to the causation of one condition by another, and clusters may fall into more than one of these categories [10]. The commonly occurring clusters identified here were a mixture of these types: shared high prevalence rates predominantly explaining the metabolic syndrome, CVD and age-related clusters; shared risk factors for cancers and renal failure; causation of subsequent conditions for clusters with mental health disorders, chronic pain and HIV/AIDS. Further research is required to delineate the causal pathways and also to enable the prediction of subsequent chronic conditions. Regardless of the category of clustering, the high levels of association between chronic conditions should inform healthcare redesign with a cluster-based and multiple-condition approach.

The clusters identified may vary according to the method employed such as the strength of clustering and the frequency or size of the cluster. However, in our analysis there was a high level of concordance between the three methods employed. There is no fixed methodology for defining clusters and it is an area that requires further research, including the methodology of identifying clusters and of their categorisation into concordant and discordant clusters.

### Healthcare Utilisation

Overall, inpatient services account for the highest increase in health spending with a 22-fold increase between having 2 and 11 conditions whilst increments in other costs such as outpatient services and medications were much lower at between 4-to 6-fold. This emphasises the need for improved healthcare delivery to achieve greater chronic disease control and secondary prevention, as being key to cost containment in MCCs.

The value of healthcare spending per patient and patterns of utilisation vary greatly by cluster regarding relative spending on medical versus pharmaceutical costs, but less so between the medical services of primary, specialist, inpatient, psychiatric and emergency care. The largest single category of healthcare spending was for branded drugs in patients with the HIV/AIDS cluster. Mental health clusters accounted for the second and third highest healthcare spending, higher than the CVD, cancer and renal failure clusters, and with the greatest spending, approximately half, on outpatient services.

The decrement of relative spending in primary care with each additional condition accrued suggests health system changes that enable MCC patients to be managed for longer in primary care could be hugely cost-saving. Spending on psychiatric services was low in all clusters, even in those clusters in which mental health disorders are present in over 95% of patients, suggesting underutilisation of psychiatric specialist services in those who could benefit. The high overall health spending in mental health clusters may reflect poorer management of additional chronic conditions [10]. Investment of resources to ensure that such patients have adequate access to healthcare for their mental health disorders is necessary to improve both the health and cost burden in such patients.

### Costs in previous literature

There is considerable variation in the magnitude of resource utilization reported between studies, health systems and data sources. Our findings align with existing evidence that MCC patients experience more complex inpatient and outpatient care scenarios leading to disproportionately high use of specialist services, visits to a multitude of physicians and confronting physicians with more problems at each visit [6,7]. MCCs patients have been reported as having more prescription medications (polypharmacy) and higher prescription drug expenditures [8,9]. However, our findings suggest that, other than for certain clusters, high utilisation of medical services is the principle factor in the elevated MCC cost burden.

Patient factors previously reported to determine cost and healthcare utilization have included age, living arrangements (e.g., living alone), being female and having supplementary insurance [24-27]. Our study shows that they may also be influenced by the number and clustering of chronic conditions.

### Demographic Variation

Age was not indicative of cost for all clusters; this finding for CVD and renal failure in particular are noteworthy and strengthen the case for primary prevention with a view to compression of morbidity to older ages.

Our study did not investigate variation by socio-economic status at individual level. Previous studies have shown in most countries a strong, negative relationship between SES and MCC among adults under 55 years but no consistently for adults older than 55 years [28]. Inequalities in access to CVD medications have been shown both between countries and by income status within countries [29].

### Implications for Health Policy

Clusters are highly amenable to large improvements in health and cost outcomes through relatively simple shifts in healthcare delivery such as the use of joint disease guidelines that tackle more than one common condition in a cluster, tailored screening and prevention. Healthcare payment mechanisms in developed countries often reward activity rather than desirable outcomes; shifting towards payment for quality or outcomes would facilitate better management of MCCs.

The variable clustering of certain chronic conditions more than others warrants urgent and careful consideration in light of the strength of such associations and the potential to have considerable impact through relatively small shifts in healthcare delivery. Mental health disorders may warrant particular attention through further recognition, prevention and screening practices, and disease management. The relatively recent phenomenon of co-existence and clustering of chronic communicable conditions with highly-prevalent NCDs such as diabetes and CVD, represents a serious threat for a failure of management of these conditions and increase in their prevalence, further complicated by poor healthcare access. Learnings from the successful delivery of HIV programs may be relevant to develop multiple disease frameworks, such as integrated care for NCDs and HIV in Kenya [30] and medication adherence clubs [31].

As many of the most important clusters identified, metabolic syndrome, CVD, mental health issues and cancers, are highly amenable to modification, greater emphasis should be placed on the role of primary prevention and lifestyle behaviour change to avoid the predicted rise in MCCs [32-34]. A study in India reported MCC rates to be highest in adults with the risk factors of alcohol (12.3%), overweight (14.1%) and central obesity (17.1%) [23].

Future work on healthcare delivery towards MCC should address its many challenges of disease burden, functional health, quality of life and healthcare costs, as well as issues related to polypharmacy.

## Limitations

Although the data used in this study are US based,, the relative changes in cost and contributors to costs would be similar in most other health systems as these are determined by the interaction and natural history of the diseases and are not dependent on the healthcare system. Other findings, such as the top ranking clusters or absolute healthcare spending, may be less generalisable to other health systems.

Certain chronic communicable conditions, such as TB, form important clusters in LMIC [35] but without sufficient prevalence in the US for TB, were not a focus of this study with the exception of HIV/AIDS.

The costs reported in this study are of total healthcare spending as it would not be feasible to distinguish costs accrued specifically from episodes directly related to chronic conditions. In addition, only costs towards healthcare appearing in financial claims were included such that other costs were not reported eg. out of pocket expenses (OOPE) that have been reported to also increase for MCC patients [36].

There is no agreed taxonomy for MCCs leading to heterogeneity in the number of conditions included and whether those include symptoms and risk factors in addition to disease end-points [10-13]. Furthermore, the interaction between clusters of conditions, for example concordant versus discordant clustering, is also important to study for purposes of prediction and prevention of subsequent chronic conditions [10].

## Conclusion

In one of the most comprehensive studies to investigate MCCs, we have reported that when applied to the US population, over half of the adult insured population have MCCs, identified the most important clusters and quantified the healthcare spending for MCCs and clusters, in a representative US patient sample. We identified that inpatient care accounts for the highest proportion of the increased spending overall but that utilisation varies greatly by clusters, which is more predictive than other patient factors. Specific healthcare interventions for MCCs should take into account the local disease burden with regards to clusters. The findings emphasis the need in any long term strategy to focus on primary prevention as the majority of the top clusters are amenable to prevention through lifestyle behaviour change. In the short and medium term, health systems should focus on secondary prevention and disease control to reduce inpatient admissions. Greater reliance on specialist care may be necessary due to the greater complexity of care, however this is inefficient whilst delivered vertically for individual conditions when one in three adults have more than one chronic condition. The goal would be the delivery of care with a multi-disease framework rather than one condition at a time, in primary or secondary care. Examples of this are emerging in developing countries for HIV and CVD [30,31,37].

Interventions for MCCs with proven health and cost outcomes are lacking. Certain interventions have started to show early impact, including the use of fixed dose combination pills to improve medication adherence and tackle undertreatment [38], cross-condition and symptom-based management guidelines, and community models of healthcare delivery [39-41]. Additional research is required to identify which interventions are impactful. Future chronic disease prevention and control approaches should be broad and patient-centric, taking into consideration healthcare payment mechanisms, the use of digital techonology, tools to help with medication use and interventions to achieve positive lifestyle change, in order to avert the alarming projections of increases in MCCs rates.

## Data Availability

The study used data from the Marketscan database, available to license through Truven Health Analytics

## Author Contributions

CH designed the study, oversaw the data analysis and wrote the manuscript. YS conducted the data analysis, assisted with study design and provided input to the manuscript. AW reviewed and provided input to the manuscript.

## Data Sharing

The study used data from the Marketscan database, available to license through Truven Health Analytics [13].

